# The Role of Prehabilitation in Improving Brain Health and Cognition After Chemotherapy in Patients with Colorectal Cancer: Study Protocol of the Chemo Brain Prehab Project

**DOI:** 10.64898/2026.01.18.26344361

**Authors:** Katie L. Hoad, Chan Ton, Deborah Williamson, Daren A. Subar, Helen E. Nuttall, Christopher J. Gaffney

**Author notes:** Corresponding Author: Dr Katie Hoad, Lancaster Medical School, Health Innovation One, Sir John Fisher Drive, Lancaster University, Lancaster, UK.

## Abstract

**Introduction:** Regional disparities in the incidence of colorectal cancer remain a concern within the UK, particularly in North West England, with 37% higher incidences than the national average. Given the growing burden of treatment-related side effects such as cognitive impairment, this study aims to investigate whether prehabilitation can improve brain health and cognitive function in patients with colorectal cancer receiving adjuvant and neoadjuvant chemotherapy.

**Methods:** This randomised control trial will recruit eighty-six patients with stage II and III colorectal cancer, who will receive adjuvant and neoadjuvant chemotherapy (fluorouracil, capecitabine, or oxaliplatin) and will be randomised between prehabilitation and standard care groups. The prehabilitation group will be provided an individualised, home-based exercise programme before and during chemotherapy, multivitamin supplementation, telephone check-ins, and activity devices. The standard care group will be provided with information about physical activity and nutrition at the start of the intervention. Habitual physical activity will be tracked in all patients. Assessments will be conducted at baseline, 72 hours before the first chemotherapy administration, and 72-96 hours after the final treatment. Outcome measures will include cardiopulmonary fitness, neurotrophic biomarkers, electroencephalographic activity, cognitive function tests, and a cognitive-related quality of life assessment. This protocol was approved by the East Midlands - Derby Research Ethics Committee, United Kingdom (approval number: 25/EM/0250) on December 17^th^, 2025. Recruitment will start early 2026.

**Discussion:** This study protocol is designed to test whether home-based prehabilitation can improve brain health and reduce chemotherapy-related cognitive impairments in patients with colorectal cancer. Insights from this work may support the development of more accessible and effective prehabilitation programmes to address treatment specific cognitive impairment.

**Trial Registration:** ClinicalTrials.gov ID: NCT07341217 (registered on January 13^th^, 2026).

## Introduction

Colorectal cancer is the fourth most common cancer in the UK, accounting for approximately 11% of all cancer cases [UK Parliament. House of Commons Library, 2024]. The incidence of colorectal cancer in the North West of England is 37% higher than the national average [NWCR, 2024], and overall survival in this region is the lowest in England at 58% (95% CI, 57 - 59%) [Franklyn et al., 2023]. Over half (54%) of colorectal cancer cases are attributed to modifiable risk factors, including tobacco use, obesity, and physical inactivity. Indeed, physical activity alone contributes to approximately 5% of all cases [Brown et al. 2018]. Most patients with colorectal cancer receive chemotherapy, with 82% in stage III cases [Upadhyay et al., 2015].

Prehabilitation could help many patients across the region each year by mitigating chemotherapy-related side effects, particularly side effects that affect cognitive function and brain health.

Prehabilitation aims to improve patient outcomes by preparing patients for the physiological and psychological demands of cancer treatments such as chemotherapy. Prehabilitation programmes are delivered as uni- or multimodal interventions, incorporating exercise, nutritional, and psychological support [Lambert et al., 2021]. Exercise-based prehabilitation can reduce the physiological and psychological side effects related to both disease and treatment, including fatigue, depression, and incontinence [Zimmer et al., 2016]. Where exercise-based prehabilitation had a positive impact on these side effects, it concomitantly improved quality of life [Mishra et al., 2012].

Another commonly reported chemotherapy-related side effect is cognitive impairment, also known as chemo brain. Reduced cognitive function is a commonly reported side effect of cancer and chemotherapy [Whittaker er al., 2022]. Specifically, patients with cancer reported and exhibited difficulties in learning, memory, attention, processing speed, and executive function [Ahles et al., 2012; Hwang et al., 2021]. Up to 85% of patients undergoing cancer treatment reported mild-to-severe cognitive difficulties, which persist for months or even years following completion of cancer treatment [Campbell et al., 2020]. In addition to significantly reduced quality of life, chemotherapy-related cognitive impairment adversely affects activities of daily living and interpersonal relationships [Falleti et al., 2006; Boykoff et al., 2009; Nuttall et al., 2025].

Based on growing evidence supporting the positive role of aerobic exercise in improving brain health in older adults with mild cognitive impairment [Farhani et al., 2022], and that regular physical activity is associated with reduced risk for vascular cognitive impairment, particularly vascular dementia in older adults [Vítor et al., 2023], exercise has emerged as a promising yet underexplored approach for managing chemotherapy-related cognitive impairment. Many cancer survivors self-report cognitive difficulties that are not fully reflected in objective measure cognitive function through test batteries [Hutchinson et al., 2012; Pullens et al., 2010]. These subjective concerns are clinically relevant, as they reflect perceived cognitive challenges in daily life [Hutchinson et al., 2012; Bender et al., 2008]. Such concerns are often associated with fatigue, anxiety, and depression [Hutchinson et al., 2012]. There remains a need for objective measures to understand the underlying mechanisms and extent of cognitive changes associated with chemotherapy.

Exercise-based prehabilitation can improve both physical and brain health. Physiologically, it improves cardiovascular function, supporting weight loss and stimulates muscle protein synthesis and skeletal muscle accretion [Lambert et al., 2021; Orange et al., 2021]. Neurologically, exercise increases circulation of neurotropic factors that support brain health and cognitive reserve [Oosterhuis et al, 2023], including brain-derived neurotrophic factor (BDNF) and vascular endothelial growth factor (VEGF) [Rich et al., 2017; Nuttall et al., 2025]. Acute aerobic exercise has been shown to increased expression of BDNF [Szuhany et al., 2015] and VEGF [Almodovar et al., 2009]. These exercise-induced molecular changes promote neurogenesis, resulting in increased gray and white matter volumes and enhanced cerebral neural activity [El-Sayes et al., 2018]. Despite these established effects of exercise, little is known about the biological mechanisms associated with prehabilitation and how they might impact cognitive function in patients with colorectal cancer. The prehabilitation is therefore hypothesised to strengthen neurophysiological resilience before chemotherapy, potentially reducing the impact of the chemotherapy-related cognitive impairment.

The overarching aim of this study is to determine whether prehabilitation before chemotherapy improves brain health and cognitive-related quality of life. A randomised controlled trial will be conducted to examine the effects of an exercise and nutrition-based prehabilitation intervention on cognitive function in patients with colorectal cancer. All patients with colorectal cancer will have their physical activity monitored throughout the study, and will undergo brain activity measurements, cardiopulmonary exercise testing, cognitive function testing, and evaluations of cognitive-related quality of life. Blood samples will be collected to assess changes in blood-based markers of neuroplasticity. It is hypothesised that patients who receive prehabilitation will show increased circulating concentrations in BDNF and VEGF. In parallel, prehabilitation will be associated with improved performance on cognitive function tests and reduced chemotherapy-related cognitive impairment, thereby reducing the negative impacts on cognitive-related quality of life following chemotherapy.

## Materials and Methods

### Overview of Study Design

The Chemo Brain Prehab Project will investigate how prehabilitation, delivered before and during chemotherapy, affects brain activity, cognitive function, neurotropic factors, cardiopulmonary fitness, and cognitive-related quality of life three months after treatment in patients with colorectal cancer. Eighty-six patients will be randomly assigned to one of two groups: (1) an individualised exercise prehabilitation programme with nutritional supplementation, or (2) standard care (control). Patients in both groups will receive chemotherapy as part of standard care. Figure 1 shows an overview of the aims and methods of the study.

**Figure 1:**
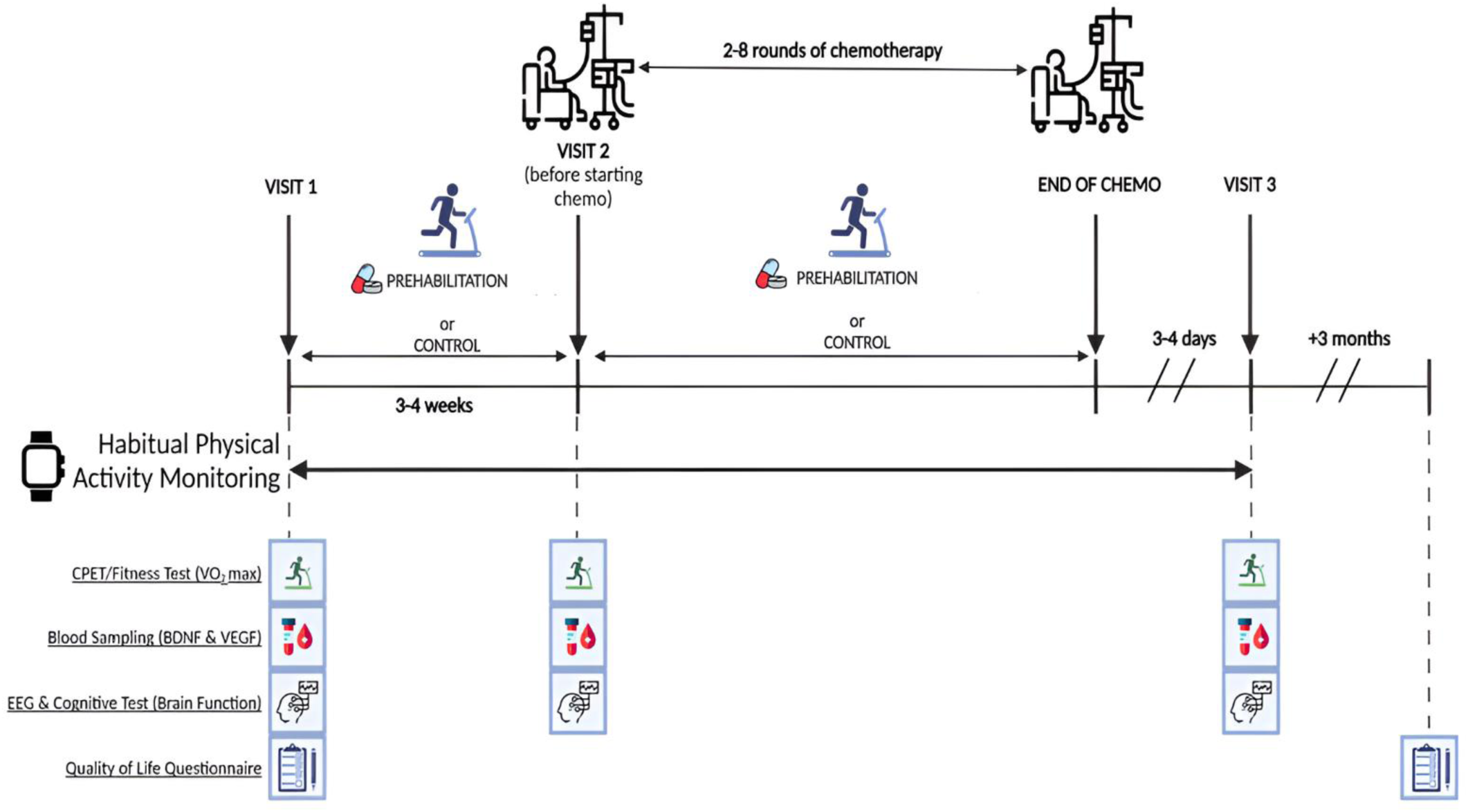
Overview of the study protocol.

#### Study Aim

This study aims to determine whether prehabilitation improves health in patients with colorectal cancer, mitigates chemotherapy relative and neural dysfunction, and whether these changes are associated with improved quality of life three months after chemotherapy. The study objectives are to evaluate whether prehabilitation improved brain health before chemotherapy by measuring physiological blood based markers of neuroplasticity, and if prehabilitation moderate the neurotoxic effects of chemotherapy by assessing changes in brain activity and cognitive function before and after chemotherapy. Additionally, the study will examine whether prehabilitation improved cognitive-related quality of life by tracking changes from baseline to three months following chemotherapy.

#### Study Setting

The study will be conducted within Lancashire and South Cumbia at East Lancashire Teaching Hospitals NHS Trust (ELTH), Lancashire Teaching Hospitals NHS Trust (LTHTR), and University Hospitals Morecambe Bay Trust (UMBHT). These sites will allow for patients with colorectal cancer to be recruited from 33 priority wards across the region [Cancer Research UK, 2022], with targeted provision in areas with the highest levels of socioeconomic deprivation and poorest health outcomes, critical in ensuring healthcare equity in prehabilitation [Stewart et al., 2025].

### Eligibility Criteria

Diagnosis of colorectal cancer is made using colonoscopy and confirmed histologically by biopsy to confirm staging of stage II or III colorectal cancer. Standard practice is to stage all patients for distant metastatic disease. For those with rectal cancer, local tumour staging is done by MRI scan or transrectal ultrasound if MRI is contraindicated.

#### Inclusion Criteria

Patients will be included if they meet all of the following criteria:

▪ Male or female, aged 60-85 years.
▪ Histologically confirmed diagnosis of stage II or III colorectal cancer.
▪ Schedules to receive adjuvant or neoadjuvant chemotherapy with fluorouracil, capecitabine, or oxaliplatin.
▪ Not engaged in structured exercise training within the six months prior to providing consent.
▪ Fluent in English.
▪ Capacity to provide informed consent.

The study will recruit equal numbers of male and female patients with colorectal cancer to enable sex-based comparisons and enhance ecological validity.

#### Exclusion Criteria

Patients will be excluded if they meet any of the following criteria:

▪ Presence of comorbidities that may alter the metabolic response to exercise (e.g., diabetes mellitus).
▪ Current musculoskeletal injury or physical limitations that prevent completion of cardiopulmonary exercise testing (CPET).
▪ Any contradiction to the multivitamin supplementation (Forceval), including known hypercalcaemia, haemochromatosis, peanut or soya allergy, current use of phenytoin or tetracycline antibiotics, and clinically significant impaired renal or hepatic function.
▪ Evidence of pre-existing cognitive impairment, including diagnosis of dementia, other neurodegenerative disorders (e.g., Alzheimer’s disease), or clinically indicated mild cognitive impairment.
▪ Diagnosis of atrial fibrillation and haematological malignancy.
▪ Participants without home internet access will be excluded due to the online delivery of the exercise programme.
▪ Diagnosis of profound hearing loss.
▪ Receiving palliative care.
▪ Presence of synchronous cancer.

### Recruitment

The lead clinician at each participating site (DS: ELHT; DW: LTHTR; CT: UHMBT) will review weekly lists of patients with colorectal cancer throughout the recruitment period, 30^th^ of January 2025 to 31^st^ of December 2025. Medical history and cancer diagnoses will be reviewed by lead clinician at each participating site to identify eligibility screening. Eligible patients will be approached during routine clinical appointments with the lead clinician and a member of the research team (KLH) and provided with an opportunity to express interest in participating.

Patients will be allowed a period of up to one week to decide on study participation if required, in which a member of the research team will contact them via telephone for their verbal consent.

Following verbal informed consent, the patient will be provided with further information about the study and, where necessary, confirm eligibility in consultation with the projects clinical lead (DS).

### Pre-Randomisation and Screening

Following verbal consent, potential participants meeting the age and diagnostic criteria will undergo detailed screening by a member of the research team via telephone (KLH). All screening questionnaires will be completed prior to the baseline assessment to confirm eligibility. Screening questionnaires will include the International Physical Activity Questionnaire (IPAQ) [Craig et al., 2003], the Physical Activity Readiness Questionnaire (PAR-Q+) [Warburton et al., 2011], Mini-Cog [Abayomi et al., 2024] and Informant Questionnaire on Cognitive Decline in the Elderly (IQCODE) [Jansen et al., 2008]. The IPAQ will be used to assess baseline activity levels and confirm the absence of structured exercise participation within the previous six months. The PAR-Q+ will be used to determine whether patients can safely engage in exercise, with medical clearance obtained if necessary. The IPAQ and PAR-Q+ will be completed via telephone. The IQCODE will be administered to screen for cognitive impairment before chemotherapy. The Mini-Cog and IQCODE will be sent through a link to the potential participants after the telephone call. The IQCODE and Mini-Cog will be administered to screen for any cognitive impairment. The Mini-Cog is a quick, 3-minute screening test for cognitive impairment for older adults, combining a 3-item word recall with a clock drawing test [Abayomi et al., 2024]. The 16 item IQCODE will be completed by the patient to assess changes in their cognitive abilities over the past 10 years and to identify mild cognitive impairment before chemotherapy [Jansen et al., 2008].

All eligible patients who are interested in taking part will be scheduled for an in-person visit for baseline assessments. At this baseline assessment visit, written informed consent will be obtained, blood samples drawn, anthropometrics, brain activity, cognitive function, CPET, and cognitive-related quality of life questionnaires (list) will be collected.

### Randomisation

A total of eighty-six patients with colorectal cancer will be randomised using a permuted block randomisation method into two groups: (1) prehabilitation (n = 43), and (2) standard care (not receiving prehabilitation) (n = 43). A computer-generated randomisation sequence will be created by the Lancaster University Health Statistics Department and stratified by sex, cancer stage, and number of planned chemotherapy cycles. The sequence will be provided to the research team in sealed, opaque envelopes.

Eligible, consented participants will be assigned a unique participant identification number and randomised on a 1:1 basis to either the exercise and nutrition-based prehabilitation group or the standard care group. The group allocation will be revealed only after baseline assessments are completed. To ensure appropriate 1:1 allocation, a designated member of the research team will review the randomisation process at the end of the recruitment period. Allocation details will remain sealed and confidential until study completion. The researcher delivering the prehabilitation intervention (KLH) will be aware of group allocation; however, all members of research team, including the clinicians named on this manuscript, will remain blinded to treatment allocation. To maintain blinding, participants will be advised not to discuss their group allocation or intervention activities with their direct care team and to direct any related queries about the study to the researcher (KLH).

### Hospital Visits

All participants will be scheduled for three visits to assess outcome measures. Visit 1 (baseline assessment), which will take place 3-4 weeks before the start of chemotherapy. Visit 2 will occur within 72 hours to the first chemotherapy infusion and Visit 3 will occur 72-96 hours after the final chemotherapy infusion.

### 3-month Follow Up

To minimise travel burden, participants will be scheduled for a telephone appointment three months following their final chemotherapy infusion. During this call, a self-reported questionnaire on cognitive-related quality of life will be administered. If a telephone appointment cannot be arranged, participants will have the option to receive the questionnaire by post for home completion.

### Intervention Group

#### Exercise Programme

Participants will be taught how to perform exercises safely at home. A qualified specialist exercise researcher (KLH) will provide exercise education and prescription to participants in the intervention. Prior to allocated to the intervention group, individualised moderate-intensity exercise programmes will be prescribed based on baseline CPET parameters and a functional fitness assessment [Jones and Rikli, 2002]. These exercise programmes will account for each participants’ physical capabilities. The intervention will be supported by the Theory of Planned Behaviour [Ajzen, 1991, Hagger et al 2022], which posits that an individual’s intention to exercise is influenced by attitudes, social norms, and perceived behavioural control. In the context of patients with colorectal cancer receiving chemotherapy, the programme will seek to foster positive attitudes towards exercise by clearly communicating the potential benefits for physical function, cognitive wellbeing, and post-treatment management (e.g., fatigue).

Subjective norms will be addressed by involving healthcare professionals and family/peers in promoting the acceptability and value of exercise as part of routine care. Perceived behaviour control will be supported through education on safe exercise practices during chemotherapy (e.g., adapting for fatigue or gastrointestinal sides effects), personalised home-based sessions, and regular check-ins to build participants’ confidence in their ability to remain active despite fluctuating treatment-related side effects. In targeting these behavioural determinants, the programme aims to encourage safe and sustained exercise engagement in physical activity throughout the course of chemotherapy.

The exercise component of the prehabilitation programme will be delivered remotely and partially supervised on a weekly basis by the qualified specialist exercise researcher. It will include four weekly sessions (approximately 40 minutes per session), comprising aerobic and resistance exercises. Each session will involve a warm up comprising of aerobic activity at 60% V̇O_2_ max, resistance exercise targeting upper and lower body muscle groups using resistance bands, followed by a cool down. Exercises will be modified through progressions or regressions based on individual capabilities. Two sessions per week will be supervised via video platform, and two will be completed independently using pre-recorded videos and an exercise programme booklet. Participants will undertake all four session per week, both prior to and during chemotherapy. Exercise tolerance will be progressively developed during visit 1 and 2, after which session frequency will be maintained throughout chemotherapy. This schedule was informed by patient and public involvement (PPI) advisors, including individuals with colorectal cancer and their carers, who considered it feasible.

#### Nutritional Intervention

At visit 1, all participants will have a blood panel that includes nutritional assessment using the Abridged Scored Patient-Generated Subjective Global Assessment (abPG-SGA) [Gabrielson et al., 2013] that will collect several markers: full blood count, urea and electrolytes, glucose, liver function tests, magnesium & phosphate, calcium and albumin, clotting function, copper, zinc, selenium, iron, ferritin, B12, folate, manganese, C-reactive protein and vitamin D. Samples will be collected and labelled before being sent to the pathology laboratory. Any abnormal nutritional biomarkers will be reviewed by the on-site dietitian as part of the participant’s standard of care. Nutritional markers will be compared between groups across the collection timepoints. Only the intervention group will be provided with an oral multivitamin supplement (Forceval Capsules, United Kingdom) by the on-site dietitian at each participating hospital. Supplements will be taken daily for a 3-4 week period before chemotherapy (between visit 1 and 2) and continued throughout chemotherapy (from visit 2 to 3).

#### Adherence To The Intervention

Adherence to the intervention will be assessed using three methods: (1) attendance record from supervised sessions (2) telephone check-ins every 1-2 weeks, and (3) data collection from wrist-worn accelerometers, Xiaomi Smart Band 9 (Xiaomi, China) to capture exercise duration and intensity during exercise sessions.

Multivitamin compliance will be assessed by requesting participants to return used capsule trays at visit 2 and visit 3.

### Standard Care Group (Control)

All participants will be seen in the outpatient setting at time points determined by each participant’s direct care team. Study visits will not interfere with any individual’s cancer treatment plan and, where possible will be aligned with routine oncology appointments to minimise travel burden.

Participants in the standard care group (control) will undergo assessments at the same time points as the intervention group. At baseline assessments (visit 1), they will receive a standard National Health Service (NHS) educational leaflet outlining physical activity and nutrition guidelines for older adults, instead of an individualised exercise programme.

Nutritional blood samples will be collected, and any abnormal nutritional biomarkers, as described above, will be reviewed by the site dietitian as part of standard care. Participants in the standard care group will not receive multivitamin supplementation that is part of the prehabilitation intervention.

To monitor habitual physical activity across groups, participants in the standard group (and the prehabilitation intervention group) will be provided with a wrist worn accelerometer (ActiGraph wGT3X-BT1, Ametris, Florida) for continuous wear.

### Outcome Measures

#### Cardiopulmonary Fitness

Participants will be advised to continue taking regular medication but will be instructed to avoid caffeine, alcohol, tobacco, and strenuous exercise on the day of each visit. For two hours prior to testing, participants will be required to fast, with water permitted. Comfortable activewear and appropriate footwear will be required.

The CPET will be conducted using a cycle ergometer, which will allow for precise determination of work rate and evaluation of the V̇O_2_-work rate relationship. Dynamic measurements include blood pressure, gaseous exchange, heart rate, electrocardiogram (ECG), and oxygen saturation. Measurement of these values will be integrated and expressed in graphical form from which other values (anaerobic threshold and V̇O_2_ max) can be derived. Gas exchange will be measured using a calibrated gas analyser, and continuous monitoring of oxygen saturation will be carried out using a pulse oximeter.

Following explanation of the protocol, participants will undergo a resting baseline phase, followed by a resistance-free pedalling phase (unloaded cycling), and then a continuous, uniform increase in work rate (incremental phase) until volitional exhaustion lasting around 10 ± 2 minutes [Chambers and Wisely, 2019]. Pedalling cadence will be maintained between 60-80 revolutions per minute (RPM). Work rate increments will be individually tailored based on physical fitness levels, with the aim of achieving maximal effort.

#### Brain Activity and Cognitive Function Measures

Participants will be fitted with a 32 channel electroencephalography (EEG; Smarting Pro, MBrainTrain, Serbia) cap equipped with saline-based hydro-links, which will measure participants’ electrical brain activity. Participants will first complete a 5 minute resting-state EEG recording, with no task present. EEG activity will be used to assess brain function, as frequency bands of the EEG are sensitive to alterations, e.g., in spectral power, which are associated with chemotherapy-related brain changes. EEG data will also be recorded during the cognitive test battery (cognitive function measures) to explore brain-behaviour associations, with subsequent analysis focusing on frequency band oscillations, particularly within the theta (4-7 Hertz (Hz)) and alpha (8-12 Hz) bands [Jaiswal et al., 2025]. Power spectral density (PSD) will be calculated to quantify the relative contribution of each band, facilitating the identification of neural activation patterns associated to cognitive sub-domains including executive function, attention, memory, and processing speed.

Cognitive function will be assessed objectively using the National Institutes of Health (NIH) Toolbox Cognitive Domain (http://nihtoolbox.org/), which comprises a series of research-administered, computer-adaptive tests. The adaptive feature of these tests is designed to minimise practice effects and reduce floor- and ceiling effects. Tests will each generate a total score, a cognitive function composite score, a fluid cognition composite score and a crystallised cognition composite score [Weintraub et al., 2014]. Age-standardised scores from three tests will be examined to assess the relevant sub-domains: executive function and attention being assessed using the Flanker Inhibitory Control and Attention Test, memory being assessed using the List Sorting Working Memory Test, and processing speed being assessed using the Oral Symbol Digit Test. To control for potential hearing loss that could confound task performance, the Words-In-Noise Test will be used to differentiate auditory deficits from chemotherapy-related cognitive impairment. All cognitive tests will be administered by a researcher (KLH), with participants using a digital tablet supported by audio guidance. Each test is expected to take 3-7 minutes to complete. The proposed test battery, and duration of set up/testing have been approved PPI advisors, who consider it feasible and relevant to the objectives of the study.

In total, the EEG and cognitive test battery session will last approximately 50 minutes, including set-up and testing duration. Detailed descriptions of these tests have been previously published, and the battery has been validated in adults and normed in individuals of similar age (60-85 years) [Weintraub et al., 2013; 2014].

#### Subjective Cognitive Measures

Cognitive-related quality of life will be assess using the Functional Assessment of Cancer Therapy-Cognitive Function (FACT-Cog) questionnaire [Kock et al., 2023]. This is a standard cancer-specific questionnaire that was developed for subjective measures of cognition and how it impacts quality of life and may capture chemotherapy-related cognitive impairment more comprehensively compared to the European Organization for Research and Treatment of Cancer Quality of Life Questionnaire-Core 30 (EORTC QLQ-C30) [Campbell et al., 2020]. The questionnaire includes a subscale for perceived cognitive impairment, perceived cognitive abilities, impact on quality of life, and comments from others. Each item will be rated on a scale from 0 (not at all) to 4 (very much), with some items reverse scored. Changes in cognitive-related quality of life will be measured at visit 1 (baseline assessment) and at three months following the completion of chemotherapy to determine whether prehabilitation mitigates decline in cognitive-related quality of life in patients with colorectal cancer.

#### Habitual Physical Activity Measures

##### All Participants

All participants, including those in the standard care group, will have their habitual physical activity measured using the activity device, ActiGraph wGT3X-BT1.This device is a triaxial accelerometer that captures bodily acceleration at a sampling rate of 30 Hz [Alvarenga et al., 2025]. The ActiGraph wGT3X-BT1 can be worn on the wrist, waist, ankle, or thigh (ActiGraph, 2020) and provides an objective assessment of physical activity with excellent reliability, demonstrated by an intraclass correlation coefficient of ≥ 0.93 for vector magnitude [Santos-Lozano et al., 2012]. Participants will wear the device on the non-dominant wrist for seven consecutive days during three distinct periods: the first week of prehabilitation before chemotherapy, the first week of chemotherapy, and the week before the final chemotherapy cycle. Data will be processed using ActiLife software, with step counts derived using the Freedson algorithm and extracted in 10-second epochs [Alvarenga et al., 2025]. Data will be exported into a Microsoft Excel spreadsheet. The device will provide measures of total movement, moderate to vigorous physical activity (minutes), non-sedentary time (minutes), and step count (steps per day) over each 7-day period. Data from these measures will be transferred directly from the device when participants return to their participating hospital site. Habitual physical activity measures obtained from this device will be used to compare the prehabilitation and standard care groups.

##### Prehabilitation Group

The prehabilitation group will use the Xiaomi Smart Band 9 to monitor activity during the exercise programme (both supervised and unsupervised) and daily living activities. This wrist-worn device includes an accelerometer, gyroscope, six-axis motion sensor, heart rate sensor (photoplethysmography, PPG), and blood oxygen sensor [Concheiro-Moscoso et al., 2025]. Participants will wear the device on their dominant hand throughout the intervention. It provides objective quantification of total daily activity through step count (steps per day), distance (metres), standing time (minutes), and time spent in moderate to high-intensity activity (minutes) [Xiaomi Inc, 2024].

The Xiaomi devices will be connected via Bluetooth with the Mi Fitness application (Xiaomi, China). Data collection will be synchronised with the application, which can remotely export measures every two weeks as CSV files accessible exclusively to the research team [Xiaomi Inc, 2024]. The application enables data to be transmitted to the device’s cloud. The lead investigator will provide prompts during supervised sessions and check-in calls to encourage device use. If participants experience skin irritation from the silicone straps, these can be replaced with nylon straps. Any day containing null values will be treated as non-wear or device failure. The device will be collected at the final visit (visit 3).

### Biological Measures

Chronic exercise will be expected to increase levels of BDNF, which has neurotrophic and neuroprotective properties and may contribute to improvements in cognitive function. BDNF and VEGF will be quantified from blood plasma using enzyme-linked immunosorbent assay (ELISA). A member of the research team, will oversee all initial visits and will be responsible for collecting and labelling blood samples before delivery to the pathology laboratory. Blood will be drawn from the antecubital vein into a 7.5 ml S-Monovette Z tube containing a clotting activator (Sarstedt, Nümbrecht, Germany). Samples will be left to coagulate at room temperature for 30 minutes before centrifugation at 2000 RCF for 10 minutes at room temperature. Serum supernatant will be aliquoted into 0.5 mL volumes and stored at -80°C within one hour of collection. Blood samples will be used for determination of BDNF and VEGF concentrations. To control for diurnal variation, all blood samples will be collected at the same time during each visit (± 2 hours).

### Demographic, clinical, and anthropometric measures

Demographic information will be collected at baseline. This will include age, sex, ethnicity, educational attainment, marital status, and socioeconomic status. In addition, a comprehensive medication inventory will be obtained to account for potential confounding effects on cognitive function and neurotrophic biomarkers. Anthropometric measures, such as height and body mass, will be measured at each study visit using a calibrated stadiometer and digital scale, respectively. Participants will be asked to remove shoes and wear light clothing to ensure accuracy. Body mass index (BMI) will be calculated using the standard formula (body mass in kilograms divided by heights in metres squared; kg/m^2^) and used as an indicator of general body composition. Relevant clinical information will be extracted from patient medical records to characterise the cancer diagnosis and treatment history. This will include the date of colorectal cancer diagnosis, cancer stage at diagnosis (according to tumour, node, metastasis (TNM) classification, or clinical staging), and details of cancer treatments received to date, including chemotherapy regiments, and further cancer treatments (e.g., surgery and radiotherapy) where applicable.

### Data Management and Monitoring

All the data collected in this study will be reported on an electronic case report and tracked through EDGE, a local portfolio management system to report study details and recruitment which is connected to the national Central Portfolio Management System (CPMS). A trial steering committee will be established which will include the members of the research team, independent academics, and a patient and public involvement advisor. A sponsor representative will also be attending meetings to ensure oversight. The committee will convene quarterly. The committee will independently monitor the study’s progress, adherence to protocol, ethics, and regulations. It will advise on operational challenges, participant safety, data integrity, and overall study delivery, escalating issues to the sponsor or funder when needed. An external authorised individual will oversee data monitoring, ensuring quality, safety, and compliance. This individual will conduct interim analyses and recommend protocol changes or trial termination, if necessary.

### Sample Size

The planned analyses will aim to determine whether: (1) prehabilitation promotes an improvement in brain health in colorectal cancer patients prior to starting chemotherapy; (2) prehabilitation protects against cognitive and neural dysfunction, to establish if prehabilitation can reduce the negative effects on the brain following chemotherapy; and (3) brain health changes following prehabilitation lead to better cognitive-related quality of life three months after cessation of chemotherapy in patients with colorectal cancer compared to standard care.

Sample size will be calculated using G*Power for repeated measures mixed models, targeting a medium effect size (f = 0.25), power of 0.80, and alpha of 0.01. A minimum of 34 participants per group will be required to achieve adequate power. To account for an anticipated 20% attrition rate, 43 participants per group will be recruited.

### Statistical Analysis

Baseline comparability between the randomised groups will be assessed descriptively, without inferential statistical testing, in accordance with CONSORT guidelines for randomised trials [Hopewell et al., 2025]. Baseline characteristics will include the following variables:

- Age (continuous)
- Sex (categorical: male/female)
- Ethnicity (nominal)
- Physical activity level (continuous)
- Cognition status, IQCODE (ordinal)
- Cancer stage (categorical: stage II and III)
- Chemotherapy regimen (categorical)
- Anthropometrics (continuous: e.g., height and body mass)
- Comorbidity status (categorical)
- Socioeconomic status (categorical)
- Education (categorical)

Descriptive statistics will be used to summarise these variables: means and standard deviations for normally distributed continuous variables, medians and interquartile ranges for non-normally distributed variables, and frequencies with percentages for categorical variables. No statistical comparisons will be conducted on baseline data, in line with best practice for reporting in randomised controlled trials.

The flow of participants through each stage of the study (enrolment, allocation, follow up, and analysis) will be depicted in a CONSORT flow diagram. This will detail the number of participants assessed for eligibility, excluded (with reasons), randomised, receiving the allocated intervention, lost to follow up, and included in the final analysis. Reasons for exclusion or withdrawal will be documented and reported where available.

#### *Primary Outcome (*what outcomes will be measured, when and how)

The primary objective is to determine if prehabilitation promotes an improvement in brain health in colorectal cancer patients prior to starting chemotherapy. A mixed analysis of covariance (ANCOVA) will be conducted with Group (prehabilitation versus control) as the between-subjects factor and Time (Study Visit 1, Study Visit 2) as the within-subjects factor, with physiological outcomes as the dependent variables. Cancer staging will be included as a covariate. The significance threshold will be adjusted to account for multiple comparisons. Additionally, to determine if prehabilitation protects against cognitive and neural dysfunction, a mixed ANCOVA will be conducted with Group (prehabilitation versus control) as the between-subject factor and Time (Study Visit 1, Study Visit 3) as the within-subjects factor, with EEG spectral ratios and cognitive outcomes as the dependent variables. Cancer staging and number of chemotherapy cycles will be included as covariates. The significance threshold will be adjusted for multiple testing.

#### Secondary Outcomes

The secondary objective is to determine if brain health changes following prehabilitation lead to improvements in quality of life three months after cessation of chemotherapy in colorectal cancer patients. A mixed ANCOVA will be conducted with Group (prehabilitation versus control) as the between subjects’ factor and Time (Study Visit 1, three months follow up after chemotherapy cessation) as the within subjects’ factor, with quality-of-life scores on the FACT-Cog as the dependent variable. Cancer staging and number of chemotherapy cycles will be included as covariates.

### Study Design Considerations

Research examining the impact of prehabilitation on cognitive function in patients with colorectal cancer is limited, with most existing studies instead conducted in other cancer cohorts receiving chemotherapy or on post-chemotherapy interventions targeting cognitive impairment. For example, these includes studies in prostate [Fitzpatrick et al., 2012], breast [Fitzpatrick et al., 2012; Hartman et al., 2018], and mixed cancer populations [Oh et al., 2012; Adamsen et al., 2006]. However, such findings may not be directly transferable to colorectal cancer, particularly given the influence of age on cognitive trajectories. Age-related cognitive decline is a normal process that varies substantially between individuals [Deary et al., 2009]; however, mild cognitive impairment is the intermediate stage between cognitive changes of normal aging and those of dementia, with those with mild cognitive impairment at an increased risk of dementia (10-15% per year) [Geda et al., 2012]. Thus, those who are identified to have cognitive impairment (non-cancer treatment related) from the IQCODE will be excluded. Furthermore, consideration should be taken to the potential of participants having auditory impairments. To control for potential hearing loss that could confound task performance of the cognitive test battery, the Words-In-Noise Test [Searchfield et al., 2024] will be used to differentiate auditory deficits from chemotherapy-related cognitive impairment. In relation to the cognitive test battery, consideration for the NIH Toolbox to be used was for comparably to other cancer studies [Hartman et al., 2018].

This study will incorporate EEG-based brain activity recording alongside the cognitive test battery using the NIH toolbox. While event triggers cannot be integrated between the EEG system and toolbox, as the platform does not support external trigger synchronisation, this limitation will be addressed by focusing on frequency band oscillations recorded during cognitive test battery across time [Foy and Foy, 2020]. Although other cancer studies have employed custom designed presentation arrays to align to EEG responses with specific cognitive events [Swainston et al., 2021], the current design prioritises feasibility and standardised cognitive outcomes. Furthermore, the NIH toolbox was selected to enable comparison with normative population data, given the absence of a healthy (non-cancer) matched control group. This approach will support interpretation of cognitive function in patients with colorectal cancer by bench marking results against standardised, age-adjusted norms. Despite the inability to establish precise event-related potentials, EEG recordings during the cognitive test battery will provide valuable information regarding task-related brain activity. Specifically, analysis will focus on oscillatory dynamics known to reflect cognitive processes. The inclusion of EEG during the cognitive test battery, rather than in isolation, will offer a more ecologically valid representation of neural engagement during task performance.

Adherence to the exercise programme during the prehabilitation period, spanning the duration before and throughout chemotherapy, will be monitored using the wrist worn accelerometers (Xiaomi Smart Band 9). The device has been selected for its ability to capture continuous raw acceleration data, allowing for objective monitoring of physical activity across varying intervention durations without the burden on the participants to interact with the watch on data recording and uploads. This continuous monitoring will complement supervised session attendance, along with the unsupervised sessions, supporting adherence assessment for both home-based sessions. The activity data extraction can give some indication to a participant’s adherence to the exercise sessions and habitual physical activity that can help to support frequent telephone check-ins, in order to optimise engagement to the intervention.

### Data Collection and Management Considerations

Data will be collected from a newly recruited cohort of stage II and III patients with colorectal cancer across four timepoints: three hospital visits and a three month follow up. End of data collection for hospital visits is estimated to be completed by the 26^th^ of May 2027, and data collection for three month follow ups is estimated to be completed by the 26^th^ of September 2027. Standardised procedures will be used across all data collection domains to ensure consistency and data quality. Data will be securely stored on Lancaster University’s encrypted cloud system, with automatic backups and access restricted to authorised research team members using university-approved computers and devices.

Data management plans will follow institutional policies, using standard file formats to ensure compatibility. Metadata standards and data dictionaries will be developed to describe variables, formats, collection methods, and coding schemes. All documentation will follow best practice guidelines to support transparency and reproducibility.

Participants will be informed that their data will be used solely for this study and will not be shared externally. Consent materials will clearly state that no secondary use of data will occur. All data handling will comply with data protection regulations and university policy, with access limited to the research team.

### Ethics Approval

Regulatory approval for this study was granted by the Health Research Authority on the 17^th^ of December 2025 (REC Reference: 25/EM/0250). The research protocol was pre-registered on ClinicalTrials.gov (https://clinicaltrials.gov/), and the NCT number is NCT07341217. The current protocol version is 1.0. The trial is sponsored by Lancaster University. All testing will conform to the declaration of Helsinki and Good Clinical Practice.

Written informed consent will be obtained from all participants before baseline testing by the lead investigator (KH) in the study. Participants will be provided with contact details for institutional leads at Lancaster University and participating sites in the participant information sheet should they have any concerns about the study. Any subsequent amendments to the protocol will be submitted for further approval to the East Midlands (Derby) Research Ethics Committee. The sponsor, Lancaster University, is responsible for authorising the amendment and completing the declaration. If a participant discloses information indicating risk to themselves or others, the NHS (publicly funded healthcare system of the UK) safeguarding procedures will be followed in accordance with National Health Service and institutional guidelines. The chief investigator (CG) or sponsor is responsible for reporting serious adverse events to the East Midlands (Derby) Research Ethics Committee that provided favourable ethical opinion for the study. Reports should be submitted using the HRA safety report form (https://www.hra.nhs.uk).

### Safety Considerations

The CPET will carry a small but recognised risk of adverse events, including exertional arrhythmias, cardiac events, or syncope [Glaab and Taube, 2022]. To minimise these risks, participants will be pre-screened using the PAR-Q+ [Warburton et al., 2011]. Where required, medical clearance will be obtained before participation. A resting electrocardiogram (ECG) may be performed before exercise testing. All CPET will be supervised by personnel with current first aid certification. Access to medical personnel and an emergency response plan will be available at each participating site. Functional movement screening will be conducted before testing to reduce the risk of musculoskeletal injury, and a graded exercise protocol with appropriate warm-up and cool-down periods will be implemented [Levett et al., 2018].

EEG recordings will involve minimal risk. However, participants may experience localised skin irritation or allergic reactions to disinfectant wipes used to clean the forehead prior to EEG cap placement. Given that participants with colorectal cancer may have increased skin sensitivity following chemotherapy, the procedure will be conducted using a fitted hydro-link cap under sterile conditions. While the EEG data will not be analysed in real time and will hold no diagnostic value, any incidental findings suggestive of clinically relevant abnormalities will be reported to the participant’s general practitioner and direct care team, following standard protocol and with the participant’s consent.

Cognitive test battery and physical assessments may cause mild mental fatigue, frustration, or emotional discomfort. The protocol will be delivered with close attention to participant well-being, and assessments will not interfere with or alter the standard of care received during cancer treatment. Participants will be advised to contact the research team should any adverse symptoms arise during or after participation.

Venepuncture for blood sample collection to assess neurotrophic biomarkers will carry minor risks, such as bruising, haematoma, infection, or vasovagal syncope. These procedures will be performed by a trained health care professional using aseptic techniques. Post-procedural care will include applying pressure to the puncture site and monitoring for any adverse responses.

Safety during home-based exercise sessions will be prioritised through several precautionary measures. Participants will be instructed to have a family member or friend present in the household during each exercise session to provide immediate support if needed. Prior to commencing the home-based component, participants will receive structured instructions on how to perform each exercise safely, with particular emphasis on posture, intensity, and pacing. Education will also be provided on recognising chemotherapy-related symptoms, such as diarrhoea, fatigue, or dizziness, which may require modification or temporary cessation of exercise. Participants will be advised to refrain from exercising during episodes of acute treatment-related symptoms (e.g. if they did not receive chemotherapy that week due to a low blood count) to protect their wellbeing. Ongoing monitoring and support will be provided to reinforce safe practice and ensure adherence to individualised exercise recommendations.

Regarding the use of multivitamins (Forceval), the principal investigator during the screening process will exclude participants with any contraindications to taking these multivitamins, can be indicated through markers and medical history on patient medical records. Routine blood count checks conducted as part of the standard of care will provide additional monitoring for potential adverse changes. The multivitamin used will be dispensed and recorded by the participating hospital, ensuring that the direct care team (oncologists, oncology nurses, and principal investigators) is aware of supplementation. Awareness of multivitamin use alongside blood count results may help the direct care team identify potential concerns determine whether further steps are needed. Participation may be terminated if clinically indicated to safeguard patient wellbeing.

Pre-screening measures will include the IQCODE and Mini-Cog [Abayomi et al., 2024; Jansen et al., 2008]. This questionnaire will be used to assess eligibility but also indicative to cognitive impairment. If responses indicate to significant cognitive impairment, the participant will be advised to consult with their general practitioner or direct care team. The study team will not provide clinical diagnoses.

### Patient and Public Involvement

Six patient and public advisors, including four individuals with lived experience of colorectal cancer (including chemotherapy and cognitive difficulties such as chemo brain) and two carers of individuals undergoing chemotherapy, will contribute across all stages of the study. These advisors bring valuable insight into the patient and caregiver experience, supporting the development of a patient-centred intervention and study design [Wareing et al., 2025]. Initial involvement focused on informing the study protocol including reviewing all patient facing materials (participant information sheet and consent forms). Advisors commented on the feasibility, acceptability, and accessibility of study testing, such as the cognitive function measures. Advisors were invited to a workshop to understand use of EEG and the battery of cognitive function, which they were able to give feedback on the testing protocol. Public advisers will be consulted on the interpretation of findings and development of dissemination strategies, including the final report and communication of results in formats appropriate for diverse audiences.

## Discussion

### Limitations

Participants in the prehabilitation group are likely to experience better outcomes partly due to more frequent interactions with healthcare professionals during the study. Evidence suggests that participants in clinical studies often report more positive care experiences and may receive additional support because of their involvement [Boaz et al., 2015]. In this study, the intervention group receives closer monitoring and regular contact through the exercise programme and check-ins, which may facilitate earlier identification and management of medical issues compared to the control group. Although both groups attend the same hospital assessment visits, this difference in ongoing interaction could introduce bias, potentially influence clinical outcomes, and confound the true effect of the intervention.

A limitation of this study is the variability in treatment duration and timing of chemotherapy among participants with colorectal cancer that may impact the generalisability neurotropic biomarkers and chemotherapy-related cognitive impairment. Unlike structured prehabilitation protocols before surgery [e.g., Pesce et al., 2024 and Lambert et al., 2024] or post-treatment rehabilitation [e.g., Hartman et al., 2018], this study does not impose a fixed duration of prehabilitation during chemotherapy. While this reflects the heterogeneity of real-world clinical care it also introduces analytical challenges. To moderate for duration and timing of treatment, advanced matching and stratification will be employed to control for differences in treatment exposure and duration.

A key consideration in this study is the impact of chemotherapy-related side effects, excluding cognitive impairment, on participants’ ability to consistently engage in the exercise programme. This could be fatigue, nausea, diarrhoea, neutropenia, and low haemoglobin levels. While the programme allows for individualised and flexible participation, some individuals may be unable to complete sessions due to temporary or prolonged treatment-related symptoms. This may introduce variability in adherence that is not reflective of motivation or acceptability, but of medical necessity. Consequently, this may bias intervention effects by reducing the overall exercise dose received or by skewing results towards participants who experience fewer or milder side effects. It is important to consider this when interpreting outcomes related to adherence and generalisability.

## Dissemination

Statistical analysis and reporting of results are expected to be completed by the end of 2027. Dissemination of study findings will occur through academic publications and conference presentations. A lay summary will be developed in collaboration with public advisors and shared with participants and stakeholders to support knowledge translation and promote equitable access to prehabilitation.

## Conclusion

In conclusion, this study will provide results on cardiopulmonary fitness, neurotrophic biomarkers, brain activity and cognitive function, and cognitive-related quality of life in patients with colorectal cancer undergoing chemotherapy. The randomised control trial will provide insights that can support the development of more accessible and effective prehabilitation programmes to address treatment specific cognitive impairment.

## Authors’ contributions

All authors contributed equally to this work.

## Data Availability

No datasets were generated or analysed during the current study. All relevant data from this study will be made available upon study completion.

## Acknowledgements

This research is funded by North West Cancer Research (NWCR, grant reference: AR2024.07GAFFNEY), and sponsored by Lancaster University. The funders and sponsor will have no role in the study design, data collection and analysis, decision to publish, or dissemination.

